# Adverse event reviews in healthcare: What matters to patients and their family? A qualitative study exploring the perspective of patients and family

**DOI:** 10.1101/2021.12.10.21267585

**Authors:** Jean McQueen, Kyle Gibson, Moira Manson, Morag Francis

## Abstract

**Objectives:** Explore what ‘good’ patient and family involvement in healthcare adverse event reviews may involve.

**Design:** Data was collected using semi-structured telephone interviews. Interview transcripts were analysed using an inductive thematic approach.

**Setting:** NHS Scotland.

**Participants:** 19 interviews were conducted with patients who had experienced an adverse event during the provision of their healthcare, or their family member.

**Results:** Four key themes were derived from these interviews: trauma, communication, learning and litigation.

**Conclusions:** Findings suggest there are many advantages of actively involving patients and their families in adverse event reviews. An open, collaborative, person-centred approach which listens to, and involves, patients and their families is perceived to lead to improved outcomes for all. For the patient and their family, it can help with reconciliation following a traumatic event and help restore their faith in the healthcare system. For the health service, listening and involving people will likely enhance learning with subsequent improvements in healthcare provision with reduction in risk of similar events occurring for other patients. Communicating in a compassionate manner could also decrease litigation claims following an adverse event. Overall, having personalised conversations and a streamlined review process, with open engagement to enhance learning, was important to most participants in this study.

## Introduction

Patients and consumers of health care should be at the very centre of the quest to improve patient safety. A major element of programmes designed to improve patient safety, is having the capacity and capability to capture comprehensive information on safety events, errors and near-misses so this can be used as a source of learning, and as the basis for preventive action in the future ^**1**^. Involvement of patients and families in reviews may reveal additional information (which is not currently being captured in some healthcare systems) which could enhance learning, assist with a person-centred approach, and support patients and families with reconciliation after adverse events ^2,3^. Organisations which represent patient voices and national enquiries highlight the lack of involvement of patients in Significant Adverse Event Reviews (SAERs) and a culture which often discounts or does not fully incorporate information highlighted by patients and families ^4,5^. When things go wrong in healthcare, patients and their families frequently have valuable information which could enhance learning for the healthcare system ^1^. They may have additional contextual knowledge, which will support the health service as they devise steps that can be taken to minimise recurrence ^6^. On the other hand, poor involvement of patients and families can lead to worsening psychological distress and increased likelihood of complaints and litigation claims ^7 8 9 10 11 12^. Whilst the NHS strives to provide safe and effective person-centred care, there is a lack of research focused on how this patient family involvement should be enacted and reflected in adverse event reviews ^13^. This study explores the perspectives and experiences of patient and family engagement during adverse event reviews in the NHS in Scotland.

### Current Practice and Research

Within NHS Scotland an adverse event is an event that could have caused (a near miss), or did result in, harm to people or groups of people ^14^. The current adverse event review process dictates that patients, service users and their families are told what went wrong, why and receive an apology for any harm that has occurred ^14^. Involvement of patients and their family varies with little detail on how best to enact person-centred engagement. In some cases, patients and families are invited to submit questions in advance of the review, whilst in others patients and families may be provided with a copy of the review findings. Less frequently are they invited to share their observations surrounding the event, what mattered (and matters) to them, and how their perspectives could enhance learning. This means their issues and concerns are not always fully known to healthcare managers, with lost opportunity to address these, and the potential for vital learning could be missed. This study aims to enhance our understanding of what ‘good’ patient and family involvement looks like from the perspectives of patients and families themselves.

### Objective

Our objective was to explore patients and families’ experience of involvement in adverse event reviews to understand what ‘good’ involvement may look like from a patient and family perspective.

## Methods

This study was explorative, using inductive thematic analysis’ techniques ^15^. Interpretative phenomenological analysis allowed exploration of individuals’ lived experience and how they make sense of this ^16^.

### Participant selection and recruitment

Participants were recruited between June and November 2021 using a variety of sources: advertisement on websites (callforparticipants.com and Care Opinion), the NHS Scotland Adverse Events Network, and a range of third sector non-government organisations.

Inclusion criteria: participant or family member experienced a serious health care incident/patient safety event in the last 10 years, resides in Scotland, are aged 18 years or over and speak English. Exclusion criteria for this study were patients and families where there was an ongoing investigation or litigation claim, adverse event did not occur within Scotland.

34 potential participants responded with interest and were sent further details by e-mail in the form of a Participant Information Sheet and Consent Form. Two participants were excluded as an adverse review was not undertaken, 4 opted not to proceed and 9 did not reply to follow up emails. A convenience sample of 19 participants provided informed consent and took part in the study. Semi-structured telephone interviews were conducted with patients or family members of patients who had experienced an adverse event in the last 10 years whilst receiving care from the NHS in Scotland.

Each participant took part in one telephone interview which was digitally recorded and subsequently transcribed. Identifying features were removed to ensure confidentiality. Participant characteristics, including the nature of the adverse event, are summarised in Table 1.

**TABLE 1:** Participant characteristics (n=19)

|  |  |
| --- | --- |
| **Gender (number of participants)** | **Female: 10**<br>**Male: 9** |
| **Age (number of participants in each age group)** | 35-44 years: 8<br>45-54 years: 4<br>>55 years: 7 |
| **Category of adverse event (number of participants in each group)** | Adult death/palliative care: 7<br>Delayed diagnosis: 1<br>Fall: 1<br>Medication error: 1<br>Mental health: 3<br>Addiction: 1<br>Suicide: 1<br>Neonatal death: 2<br>Surgical complication: 2 |
| **Duration since adverse event (number of participants in each group)** | < 1 year ago: 2<br>1-5 years ago: 13<br>5-10 years ago: 4 |
| **Patient or patient representative (number of participants in each group)** | Patient: 4<br>Patient carer or family member: 15 |
| **Employment status** | Full-time: 5<br>Unemployed: 2<br>Retired: 4<br>Full-time carer: 2<br>Unable to work due to disability: 6 |

### Research team and reflexivity

Three researchers (JM, MF, MM) who are experienced in qualitative interviewing completed the telephone interviews. JM is a Principal Educator within NHS Education for Scotland and registered occupational therapist, MF is a reviewer with Healthcare Improvement Scotland and registered nurse, MM is a senior reviewer within Healthcare Improvement Scotland. The interview guide, questions and prompts were reviewed by a patient representative and updates were made following review. JM and KG (a Critical Care doctor) independently coded the transcripts of interviews. Wider members of the research team had the opportunity to read interview transcripts and commented on cross-sectional analysis and agreement of themes.

### Ethical approval

Ethical principles were followed as outlined in the Medical Research Council’s ‘Principles and guidelines of good research practice’ ^17^. Ethical approval was obtained from the West of Scotland Research Ethics Committee. As part of this approval, each participant received a written participant information sheet, advising that participation was voluntary and assuring the person that they could decline to answer any question that they felt uncomfortable with and they were at liberty to withdraw at any time without consequence. Informed written consent was obtained before participants were enrolled in the study

### Data management and analysis

Semi-structured telephone interviews, lasting between 60 and 90 minutes, explored participants’ experience of the adverse review process and their perceptions of what ‘good’ patient and family involvement would look like. Audio recordings were transcribed verbatim and stored and analysed using Nvivo 1.5.1 software. Interpretative phenomenological analysis (IPA) allowed exploration of individuals’ lived experience and how they make sense of this ^16^. Transcripts were analysed using the ‘inductive thematic analysis’ technique described by ^15^. This six-step process involves familiarisation with the data reading and re-reading the transcript, generation of initial codes, identifying themes, refining and reviewing themes and naming themes. The transcripts were coded independently by two authors (JM and KG). New themes were added as they emerged during the subsequent analysis of transcripts. Resulting themes and the point at which data saturation became apparent were discussed. Emergent themes were then shared with the study team and agreement of final themes was reached. A preliminary report was circulated to participants and their feedback used for additional validation.

This ensured credibility and that participants recognised and accepted the themes identified in this paper. Study methods utilised the Consolidated Criteria for Reporting Qualitative Research guideline ^18^.

## Results

The four themes derived from the data analysis are highlighted in Table 2 and illustrated further by narrative quotes and discussion of congruent and diverse views amongst participants.

**Table 2:** Superordinate themes and subthemes.

| Superordinate theme | Subthemes |
| --- | --- |
| **Communication: the importance of feeling listened to and included** | Being listened to, a person-centred approach, receiving an apology, feeling included, reconciliation |
| **Trauma: the challenges experienced during the review process** | Review processes were lengthy, frustrating, exhausting, negative effect on mental health |
| **Learning: the importance of demonstrating change, and improving the healthcare system and patient safety** | Closing the loop, systems thinking, addressing safety and how to improve the system or processes that contributed to the safety event |
| **Litigation: the opportunity to get answers where it was difficult to obtain answers elsewhere** | Getting answers, assurance, litigation being a last resort where answers were not obtained elsewhere |

### Theme 1: Communication - the importance of feeling listened to and included

Being listened to, feeling heard and having a person-centred approach where people felt included was important for participants during review processes. The style and method of communication and asking what really mattered to that person or their family was highly valued. There was direct contrast between those who perceived the communication personable with those who felt the communication style didn’t consider their needs and preferences. Use of the word ‘statistic’ and the focus on provision of a leaflet as opposed to dialogue in the excerpt below indicates lack of person-centred approach in some review experiences:

> *‘The lack of communication led us to feel like a statistic rather than a person. It was such an impersonal approach’* (participant 4).

Participants spoke about not being given the opportunity to discuss their individual circumstances and what happened. Instead, they were given a procedurally focused approach such as being issued with a leaflet or other type of standardised response echoed in the next excerpt below:

> *‘she said “we’ve decided that we’re going to do…a serious adverse incident review…and that I’m going to send you a leaflet”; no communication, no time to explain, we’ll just send you a leaflet… I’ve just lost my son…*.. *we’ll send you a leaflet, it didn’t feel helpful at all*’ (participant 8).

The human side of communication, asking about peoples’ preferences, including them in the process with timely person-centred dialogue, was overwhelmingly important. The extract below demonstrates how the lack of this led to feelings of helplessness, frustration and even anger:

> *‘I was never asked about what mattered to me or what type of method of communication worked best. If they had, they’d have known I wasn’t interested in the serious adverse event review, their long-winded report, or monetary compensation, I just wanted answers and to move on’*. (participant 18)

An excerpt from another participant adds to the perspective on how involving patients or their family in the review could help to support learning:

> *‘Perhaps have a bit more thought about how families should be engaged with might only need a short conversation, is there anything we need to know? Anything over and above what we have gathered that we [the NHS] need to know? They would have been able to gather from us very quickly that these are the key risks. I think that they could have drawn a lot more information from us [family] but basically that is lost because it is all very transactional - here is the response, this is what we are doing’ (participant 12)*.

In contrast, participants who felt included and listened to felt more confident about the safety of the healthcare system and were more satisfied:

> *‘I was heard and it made me feel safe going forward in the future because I’m likely to have this (medical issue) again and I’m likely to be seen (by that healthcare professional) again…. So, it made me feel incredibly safe, it made me feel heard. And it was like, actually, that’s all I want, that’s all I need to feel safe going forward’ (participant 5)*

Speaking with and including patients and their families in a compassionate way, as illustrated in the excerpt below, helped and was almost restorative following the traumatic loss of a baby:

> *’Our communication with the consultant…was really good…*.*because she was being like a human being, a women who’s a mother herself and she kind of slightly stepped back from her professional role and just spoke to you like an adult*…*it made us feel good because we knew she cared’* (participant 17).

### Theme 2: Trauma – the challenges experienced during the review process

This theme represents the challenges patients and their families experienced with the review or process. Participants reflected on the length of time it took for the review process to be completed. In the narrative below perceived inactivity during a lengthy review could impact negatively on mental health and lead to feelings of frustration and anxiety. Not being offered answers or a timely explanation contributed to negative views and impacted on wellbeing during what were already challenging times.

> *‘We are drawing this [the review] out longer and longer and longer. And I have to be careful, I don’t drown myself in this whole process…I shouldn’t have to sacrifice my own health and wellbeing just to get answers’ (participant 8)*.

Participants spoke of the long time it sometimes took review processes and the importance of timely communication and the frustration and the hurt when timescales were missed:

> *‘I just wanted it to be over because it was quite stressful. I mean, they made a big mistake, lots of mistakes, and there wasn’t an end to it for me, it was just dragging on’ (participant 2)*.

Whilst in many cases an initial response appeared to be rapid, the subsequent provision of information was sometimes lacking and this led to frustration and submission of a complaint:

> *‘And within two weeks of putting in a complaint, I did have a meeting with the associate medical director. But after that, it seemed to me really slow and took almost two years…which is a long time to have it hanging over you. So, there was a lot of time between these meetings and letters where nothing was happening’ (participant 2)*.

One participant spoke about the lengths that they had gone to convey their experience following the death of a family member by writing a detailed letter. The lack of response led to anxiety and added to the trauma experienced:

> *‘Well, it just didn’t feel great [the NHS response]. Since then I have really thought about how the NHS responded to my letter…I had taken time over the timeline …*.*every single word I poured over and thought about because I wanted to present my situation and the things I felt wasn’t right…*.., *my language was very careful, so a week later, not to have received anything, every day I thought, when am I going to hear? I was nervous. I just wanted to hear back… my expectations were to receive something. So, to get nothing and then to have to write again it just felt like adding insult to injury’ (participant 3)*.

When timeframes were missed or extended this often led to a negative perception of the review process and additional stress and dissatisfaction:

> *‘It’s terrible…they sent an email saying that they want more time. They don’t even tell you how much more time they want, it’s frustrating. They initially offered a date…. but then, you know, that passed and no timescale of when they think they will have it looked at. It was just very much…open-ended’ (participant 1)*.

Feelings of frustration and anger resonated through many of the interviews when follow-up communication did not occur. When there was no response to questions asked this could lead to suspicions of a cover-up and led participants to wonder if the service was hiding something.

> *‘The scary bit is I am going to start laying the blame at them. And that was never the purpose of my questions. It was for my own satisfaction that I want to know that things were being done. But now I’m beginning to feel things were not done, and there was negligence going on’* (participant 8).

Overwhelmingly, how the communication happened (or in some cases did not happen), and the timeframes involved were important to participants and are reflected on in the third theme, Learning.

### Theme 3: Learning – the importance of closing the loop, and improving the healthcare system and patient safety

Closely linked to the earlier theme of communication and involvement is learning; this was important to all participants. Lack of engagement with patients and families contributed to fear of missed opportunities for improvement in the healthcare system and the same adverse event occurring to other patients. Although an apology was important, it was important to many participants that they knew what changes had been made following the adverse event:

> *‘in terms of proper engagement…*.*it would have been good to see what actually changed as a result …we don’t know, and we will never know, actually, because the complaint was closed at that point because essentially we were satisfied that the complaint was upheld’ (participant 12)*.

Again, the procedural nature of the response was spoken about with limited evidence of improvement:

> *‘They just basically ticked the box, apologised for everything, upheld everything, and then it’s like no further action. I can just file that. That’s what I kind of feel because having gone back into the hospital, I don’t particularly see that there’s been much change’ (participant 12)*.

This excerpt and the one that follows highlighted participants experience of some parts of the healthcare system which may not have prioritised learning and improvement. This was disappointing for participants, many who had experienced the loss of a loved one or significant harm themselves. The overwhelming intention was to lessen the chance of something similar happening to others. The excerpts below highlights instances where the healthcare system did not lend itself to these changes:

> *‘complaining…gets me nowhere, people shut down, notes go missing, people close ranks. And then you’re not heard, and you’re not believed and actually they put the blame on me and say, oh, no, you’re paranoid or whatever. I’ve had the whole works and also …people are only human, we’re dealing with human beings that are stressed out often’ (participant 5)*.

In these excerpts participants focus on the healthcare system, the pressure staff could be under and the importance of learning and not blaming individuals, although it was interesting that in the except below a senior leader in healthcare suggested greater individual responsibility:

> *‘I suppose there is anger with me as well, but it’s just the system is not working, it’s broken and I’m just very frustrated and I think as I said to them (the chief executive), I’m not looking to put anybody’s head on the block here. It’s a system that’s not functioning properly. It was also pointed out to me that there is individual responsibility to make the system work and if people are not taking individual responsibilities properly then it’s not going to work’ (participant 8)*.

Participants recognised the strain the healthcare system is under and the potential for human error, and, in the excerpt above, apportioning blame was not the intention, but the participant appears to suggest that in one instance a senior leader within healthcare was focused on ‘individual responsibility’ as opposed to a more system-based approach:

> *‘I wasn’t looking for anybody and I am still not. My philosophy in life is that people make mistakes, we are all human and we make mistakes. Things are not going to work unless they (the health service) listen and then implement some sort of action’ (participant 8)*.

### Theme 4: Litigation – the opportunity to get answers where it was difficult to obtain answers elsewhere

Overall, where participants did not feel included, listened to and supported in a compassionate way, or where the service did not evidence there had been learning, or there was a lack of feedback and timely communication, this increased the likelihood of seeking legal advice.

Seeking compensation was never the original intention of any of the participants, as evidenced in the excerpt below. Learning mattered more with litigation being an absolute last resort and used only when attempts to get answers and improvement had not been successful:

> *‘Right from the very beginning, people had said to go straight to a solicitor, but I didn’t want to do that. I wanted just to make sure it never happens to anybody else. However, in the end, I thought that I’ve got nowhere, I really don’t feel that they are taking much responsibility, so I just decided I would take it further*’ (participant 15).

This is echoed by another participant who states:

> *‘I went two years and nine months without ever wanting compensation, and I’ve made that very clear from day one that was never my goal and I didn’t want to profit (from the death of my loved one). But I decided to do this because I was being ignored and I knew that I’d get a reaction’ (participant 13)*.

Some participant’s narratives focused on how the lack of inclusion forced them to seek legal advice, with their perception that healthcare services appeared concerned about the potential for blame; litigation was used as a method to encourage engagement and get answers:

> *‘I just feel that the medical profession is so scared of being sued that it closes down*…*if they listened to people, and tried to rectify the mistakes, in a way that people actually wanted, there would be less compensation and it’s less confrontational’ (participant 5)*.

Within this theme, participants appear to outline how a more inclusive approach would not only be restorative for them it could be less adversarial for all involved with the potential to reduce litigation claims.

This participant recalled their personal experience with use of the word ‘scared’ indicative of how those in the health service appeared:

> *‘I’ve had medical records go missing when I put in a formal complaint. I think people are scared of being sued and don’t want to take accountability. I think the NHS is so scared of being sued and it needs to get over it actually, we need to own up, we need to own our mistakes’, actually people want less money, not more. And it takes a lot less time for the NHS than going through the courts and you’d pay the lawyers a lot less*.*’ (participant 5)*.

## Discussion

Findings from this study expand our understanding on patient and family experience and their perceptions of what ‘good’ patient and family involvement in adverse event reviews might look like. The interrelated themes depict the participants’ views on challenges with communication, lack of involvement and the importance of listening to what matters to them. During the qualitative interviews participants spoke freely on their experiences around lack of personalised communication and limited inclusion in the review process. This led to frustration and impacted on their wellbeing with some stating the only way to get answers was to force this through litigation. These findings concur with similar work in the Netherlands focused on suicide reviews ^6^ and a UK based study on parental engagement following perinatal mortality ^8^ where better inclusion of patients and families supported reconciliation, learning and reduced the likelihood of litigation ^7^. Similarly a mental welfare survey found almost two-thirds of carers and families felt their views were not sufficiently taken into account following death of a family member whilst under a compulsory treatment order ^19^.

Participants illustrated the review process was long and arduous and added to an already traumatic event. Participants suggested the following aspects, which if enacted, could make a real difference. Timely person-centred communication, early involvement inviting patients and their families to provide additional information to complement the review undertaken by healthcare professionals, with their contributions offering a further opportunity to enhance learning. Patients and their family will experience the event from a different perspective and potentially have valuable information on the systems and processes leading up to the event. Communication which focused on what matters to the patient or family should feel inclusive and not a procedural or tick box exercise. Crucial to participants satisfaction was ‘closing the loop’ (proving to patients and family that you have heard their feedback and are taking it seriously) demonstrating consideration of changes to healthcare systems and services to lessen the likelihood of recurrence with future patients. Interestingly much of this concurs and builds on previous studies ^20^ and legislative ‘Duty of Candour’ ^21^ where patients who experience harm during the provision of their healthcare are offered an explanation, an apology, and informed of changes made to prevent future incidents ^12^. Our findings suggest participants would like the opportunity to feel more engaged in adverse event reviews going forward.

A limitation of our study is that there was likely recruitment bias: most participants who responded to this study had a negative experience during adverse review processes and there may be patients and families who have had positive experiences but were less likely to share these. Nevertheless, the study provides very valuable insights and experiences which we hope will inform future improvements in adverse event review processes.

Whilst much has been achieved in the field of co-production, person-centred care, involving people in healthcare decision-making more widely, including the patient in patient safety remains an issue ^22^. That said, the impact of an adverse event differs from most other healthcare interactions. Patients have been harmed, unintentionally, by the people or healthcare system in which they placed considerable trust, so their reaction may be especially powerful. This may require particular conditions within the healthcare system and specific skills and competencies for healthcare staff. There are some examples where it is beneficial to both patients and healthcare staff of patient involvement in learning from when things go wrong in healthcare ^23^. A barrier to the openness and learning required to improve safety relates to perceptions around the healthcare system or professionals fear of being blamed, reputational damage, negative media coverage and litigation ^24, 25^. Tackling this requires the fostering of a ‘just culture’ where frontline staff feel able to explain conditions that contributed to the adverse event, and able to report mistakes within a health system focused on improvement and learning where individuals are not held accountable for system failings ^26,27^.True psychological safety perhaps requires some fundamental cultural changes if true just culture and candour are to be realised patients, families and staff. Whilst this and other publications have now documented a clear direction of travel for inclusion of patients in patient safety, the focus should now firmly be on creating the conditions for openness and learning. We suggest that this focus should be on promoting culture change, psychological safety, systems thinking, exploring opportunities for guidance, training and support for governance leads, clinicians, senior leaders and risk management staff to fully enact the personalised approach outlined by the participants in this study.

## Conclusions/Key findings

This study illustrates what matters to patients and families using their suggestions to discuss improvement in practice. It adds detail on how best to enact this inclusion within adverse event reviews. Findings suggest that an open, collaborative, person-centred approach which listens to, and supports, people following an adverse event results in better outcomes for all. For the patient or their family, it can help restore their faith in the healthcare system and reassure them that learning gained may lessen the chance of similar events happening to others. For the health service, not listening to people risks missing vital learning which could improve future patient safety and quality of care. Engaging patients and families in reviews and communicating in a compassionate manner could also decrease litigation claims. Personalised conversations, a streamlined review process, focused on the healthcare system and circumstances around the event with open engagement to enhance learning were what mattered most to our participants.

## Data Availability

All data produced in the study are available upon reasonable request to authors

## Acknowledgements

The authors are very grateful to all patients and family members who volunteered their time, shared their experiences and contributed to this greater understanding of patient and family engagement during adverse event review processes. The authors are also very grateful to those who reviewed the interview schedule and commented on earlier drafts of this manuscript.

